# Omicron infection of vaccinated individuals enhances neutralizing immunity against the Delta variant

**DOI:** 10.1101/2021.12.27.21268439

**Authors:** Khadija Khan, Farina Karim, Sandile Cele, James Emmanuel San, Gila Lustig, Houriiyah Tegally, Yuval Rosenberg, Mallory Bernstein, Yashica Ganga, Zesuliwe Jule, Kajal Reedoy, Nokuthula Ngcobo, Matilda Mazibuko, Ntombifuthi Mthabela, Zoey Mhlane, Nikiwe Mbatha, Yoliswa Miya, Jennifer Giandhari, Yajna Ramphal, Taryn Naidoo, Nithendra Manickchund, Nombulelo Magula, Salim S. Abdool Karim, Glenda Gray, Willem Hanekom, Anne von Gottberg, COMMIT-KZN Team, Ron Milo, Bernadett I. Gosnell, Richard J. Lessells, Penny L. Moore, Tulio de Oliveira, Mahomed-Yunus S. Moosa, Alex Sigal

**Affiliations:** Africa Health Research Institute, Durban, South Africa; School of Laboratory Medicine and Medical Sciences, University of KwaZulu-Natal, Durban, South Africa; KwaZulu-Natal Research Innovation and Sequencing Platform, Durban, South Africa; Centre for the AIDS Programme of Research in South Africa, Durban, South Africa; Centre for Epidemic Response and Innovation, School of Data Science and Computational Thinking, Stellenbosch University, Stellenbosch, South Africa; Department of Plant and Environmental Sciences, Weizmann Institute of Science, Rehovot, Israel; Department of Infectious Diseases, Nelson R. Mandela School of Clinical Medicine, University of KwaZulu-Natal, Durban, South Africa; Department of Internal Medicine, Nelson R. Mandela School of Medicine. University of Kwa-Zulu Natal; Department of Epidemiology, Mailman School of Public Health, Columbia University, New York, NY, United States; South African Medical Research Council, Cape Town, South Africa; Division of Infection and Immunity, University College London, London, UK; National Institute for Communicable Diseases of the National Health Laboratory Service, Johannesburg, South Africa; SAMRC Antibody Immunity Research Unit, School of Pathology, Faculty of Health Sciences, University of the Witwatersrand, Johannesburg, South Africa; Institute of Infectious Disease and Molecular Medicine, University of Cape Town, Cape Town, South Africa; Department of Global Health, University of Washington, Seattle, USA; Max Planck Institute for Infection Biology, Berlin, Germany

## Abstract

Omicron variant (B.1.1.529) infections are rapidly expanding worldwide, often in settings where the Delta variant (B.1.617.2) was dominant. We investigated whether neutralizing immunity elicited by Omicron infection would also neutralize the Delta variant and the role of prior vaccination. We enrolled 23 South African participants infected with Omicron a median of 5 days post-symptoms onset (study baseline) with a last follow-up sample taken a median of 23 days post-symptoms onset. Ten participants were breakthrough cases vaccinated with Pfizer BNT162b2 or Johnson and Johnson Ad26.CoV2.S. In vaccinated participants, neutralization of Omicron increased from a geometric mean titer (GMT) FRNT_50_ of 28 to 378 (13.7-fold). Unvaccinated participants had similar Omicron neutralization at baseline but increased from 26 to only 113 (4.4-fold) at follow-up. Delta virus neutralization increased from 129 to 790, (6.1-fold) in vaccinated but only 18 to 46 (2.5-fold, not statistically significant) in unvaccinated participants. Therefore, in Omicron infected vaccinated individuals, Delta neutralization was 2.1-fold higher at follow-up relative to Omicron. In a separate group previously infected with Delta, neutralization of Delta was 22.5-fold higher than Omicron. Based on relative neutralization levels, Omicron re-infection would be expected to be more likely than Delta in Delta infected individuals, and in Omicron infected individuals who are vaccinated. This may give Omicron an advantage over Delta which may lead to decreasing Delta infections in regions with high infection frequencies and high vaccine coverage.

---

The Omicron variant of SARS-CoV-2, first identified in November 2021 in South Africa and Botswana^1^, has been shown by us^2^ and others^3-8^ to have extensive but incomplete escape from immunity elicited by vaccines and previous infection, with boosted individuals showing better neutralization. In South Africa, Omicron infections led to a lower incidence of severe disease relative to other variants^9,10^, although this can be at least partly explained by pre-existing immunity^2^. While Omicron infections are rising steeply, many countries still have high levels of Delta variant infection. How Delta and Omicron will interact is still unclear. One possibility is that Omicron and Delta will coexist, and another is that Omicron will curtail the spread of Delta by eliciting a neutralizing immune response against Delta in Omicron convalescents, so that Delta could not effectively re-infect.

We investigated whether Omicron infection elicits neutralizing immunity to the Delta virus. We isolated Omicron virus from an infection in South Africa (see Table S1 for detailed genotypic information of the viral isolate used). We neutralized this isolate with plasma from participants enrolled during the Omicron infection wave in South Africa, with each participant having a confirmed diagnosis of SARS-CoV-2 by qPCR. To quantify neutralization, we used a live virus neutralization assay and calculated the focus reduction neutralization test (FRNT_50_) value, the inverse of the plasma dilution required for 50% neutralization, as measured by the reduction in the number of infection foci. We enrolled 25 participants late November and December 2021. Two participants had advanced HIV disease based on a low CD4 count (<50 cells/uL) and unsuppressed HIV infection. Our previous data indicated an atypical response to SARS-CoV-2 infection in advanced HIV disease^11^ and we excluded the two participants from this analysis. Table S2 summarizes the characteristics of the remaining 23 participants.

Fourteen out of 23 participants were admitted to hospital because of Covid-19 symptoms, but only one required supplemental oxygen. Ten participants were vaccinated and had a breakthrough Omicron infection. Five were vaccinated with two doses of Pfizer-BNT162b2 and 5 with Johnson and Johnson Ad26.CoV2.S, with one Ad26.CoV2.S vaccinee being boosted with a second Ad26.CoV2.S dose (Table S3). Out of the 23 participants, only 3 (1 vaccinated and 2 unvaccinated) self-reported having a previous SARS-CoV-2 infection (Table S3). Participants were sampled at enrollment, which was a median of 5 days (interquartile range 3-8 days) post-symptom onset, and again at weekly follow-up visits which were attended as practicable because of the Christmas holidays in South Africa. The last follow-up visit was a median of 23 days (interquartile range 17-25 days) post-symptom onset (Table S2). Virus from the upper respiratory tract from each participant was sampled using a combined nasopharyngeal and oropharyngeal swab, and all viruses successfully sequenced were confirmed to be Omicron (Table S3).

We analyzed neutralization at enrollment (baseline for the study) and the last follow-up visit. We observed that Omicron neutralization increased in vaccinated individuals from a low geometric mean titer (GMT) FRNT_50_ of 28 at the enrollment visit to FRNT_50_ = 378 at last follow-up, a 13.7-fold increase (95% CI 3.8-49.5, Fig 1A). The samples from unvaccinated participants neutralized at a similar starting level at study baseline (FRNT_50_ = 26) but reached a lower final level (FRNT_50_ = 113) at last follow-up, a 4.4-fold increase (95% CI 1.4-13.5, Fig 1B).

**Figure 1:**
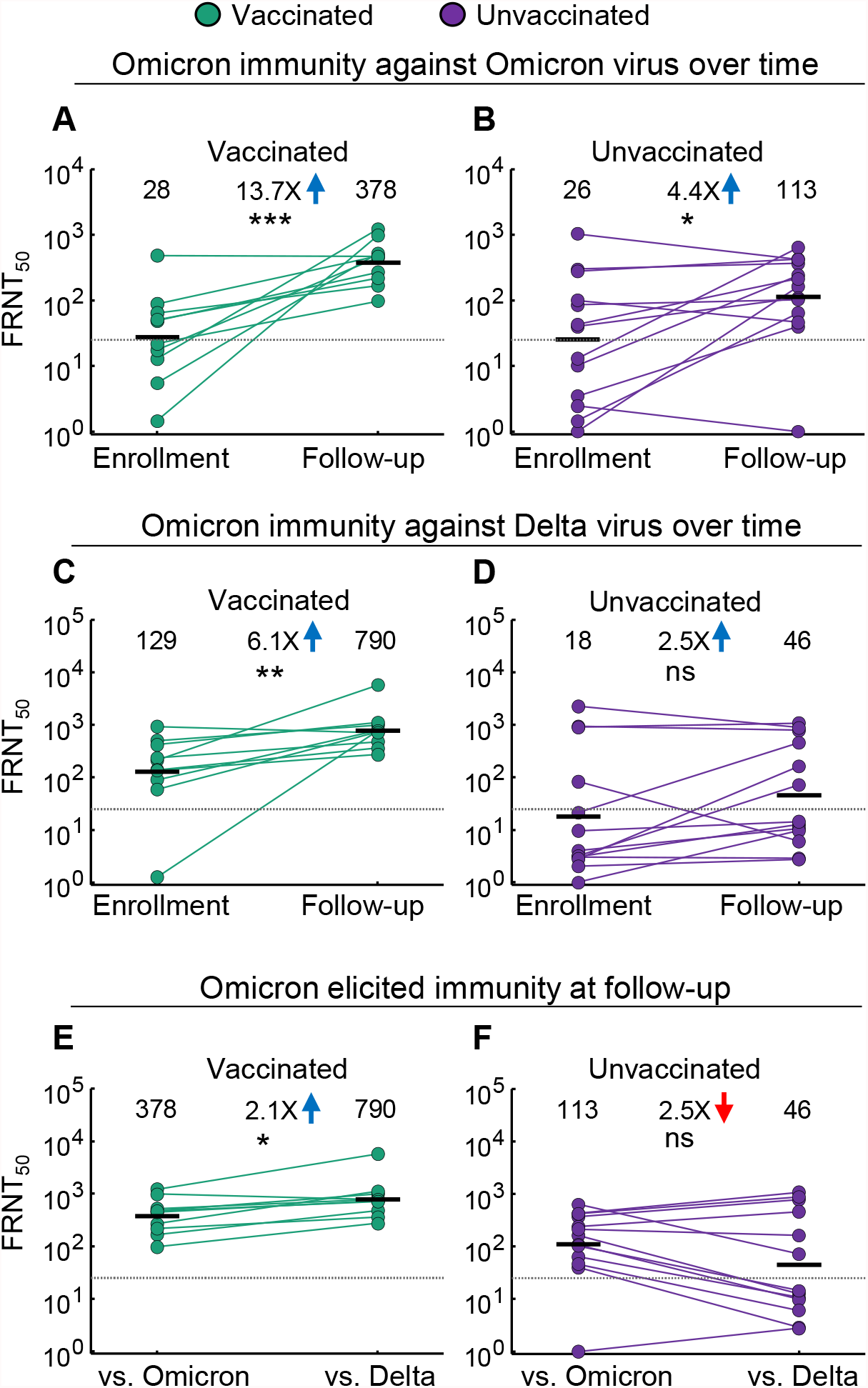
Enhancement of Delta neutralization by Omicron infection. (A) Neutralization of Omicron virus by Omicron infection elicited plasma in n=10 convalescent vaccinated participants, (n=5 two doses of Pfizer BNT162b2, n=5 Johnson and Johnson Ad26.CoV2.S). Each participant was sampled at the initial enrollment visit (median 5 days post-symptom onset) and compared to the last follow-up visit (median 23 days post-symptom onset). Numbers are geometric mean titers (GMT) of the reciprocal plasma dilution (FRNT_50_) resulting in 50% neutralization. Fold-change is calculated by dividing the larger GMT value by the smaller value and arrows indicate direction of change between enrollment and follow-up. Dashed line is most concentrated plasma tested. (B) as in (A) for the n=13 unvaccinated participants. (C) Neutralization of Delta virus by Omicron infection elicited plasma in the vaccinated participants. (D) as in (C) for the unvaccinated participants. (E) Neutralization of Omicron compared to Delta virus by Omicron infection elicited plasma in vaccinated participants at the last follow-up visit. Arrow indicates direction of change between Omicron and Delta virus. (F) as in (E) for the unvaccinated participants. p-values were: (A) 6.6 *×*10^*−*4^, (B) 0.031, (C) 2.3 *×*10^*−*3^, (D) 0.15, (E) 0.032, (F) 0.79 as determined by the Wilcoxon rank sum test.

Neutralization of Delta virus increased during this period in the vaccinated individuals. At enrollment, neutralization capacity against Delta virus was higher than against Omicron (FRNT_50_ = 129) and reached FRNT_50_ = 790 at last follow-up, a 6.1-fold increase (95% CI 1.8-20.7, Fig 1C). The unvaccinated had lower Delta neutralization at baseline with Delta virus FRNT_50_ = 18, and reached FRNT_50_ = 46, a non-statistically significant 2.5-fold increase (95% CI 0.9-7.0, Fig 1D).

Comparing Omicron and Delta neutralization at the last available follow-up visit showed that vaccinated participants were able to mount a better neutralizing response against the Delta virus than against the Omicron virus: neutralization of Delta virus was 2.1-fold higher than Omicron (Fig 1E, 95% CI 1.5-2.9). In contrast, in unvaccinated participants, neutralization of Delta was 2.5-fold lower relative to Omicron (95% CI 1.1-5.8), although this was not statistically significant (Fig 1F).

Examining neutralization at all available timepoints per study participant showed that neutralization of the Omicron variant seemed to peak approximately 2 weeks post-reported symptom onset date (Fig 2). The pattern in vaccinated individuals showed a high degree of uniformity, with a rise in Omicron neutralization capacity mirrored by a rise in Delta neutralization capacity in 9 out of 10 vaccinated participants, and with Delta neutralization level very similar to or higher than Omicron neutralization level. In contrast, the pattern in unvaccinated participants was much more variable, with neutralization of Omicron visibly stronger than neutralization of Delta virus in 6 out of 13 participants.

**Figure 2:**
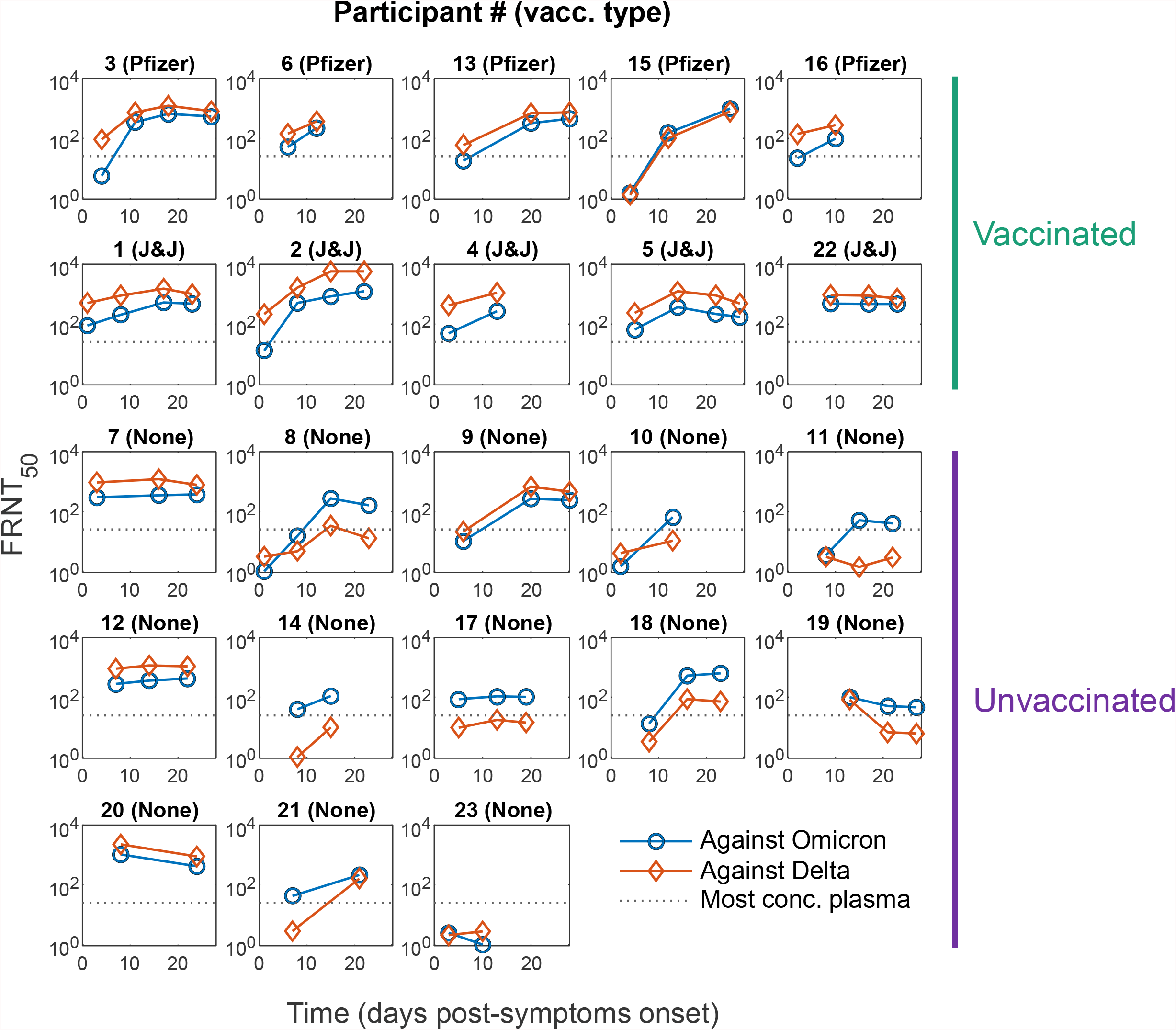
Omicron and Delta neutralization capacity over time in Omicron infected participants. Neutralization of Omicron (blue) and Delta (red) at all study visits. Participant number is as in Table S3. First row are Pfizer BNT162b2 vaccinated, second row are Johnson and Johnson Ad26.CoV2.S vaccinated, and bottom three rows are unvaccinated participants. X-axis is the time post-symptom onset when sample was collected, and y-axis is neutralization as FRNT_50_. Dashed line is the most concentrated plasma tested.

We also tested neutralization of Omicron by Delta variant elicited immunity. We collected 18 plasma samples from a group of 14 participants previously infected in the Delta variant wave in South Africa, some of whom were vaccinated either before or after infection (Table S4; for 4 of the vaccinated participants, a sample was available post-infection, and then again post-vaccination). Confirming previously reported results^7^, we observed extensive escape of the Omicron viral isolate used here from Delta elicited immunity across all samples tested. This was manifested as a 22.5-fold decrease (95% CI 14.4-35.0, Fig 3) of Omicron virus neutralization compared to Delta virus neutralization.

**Figure 3:**
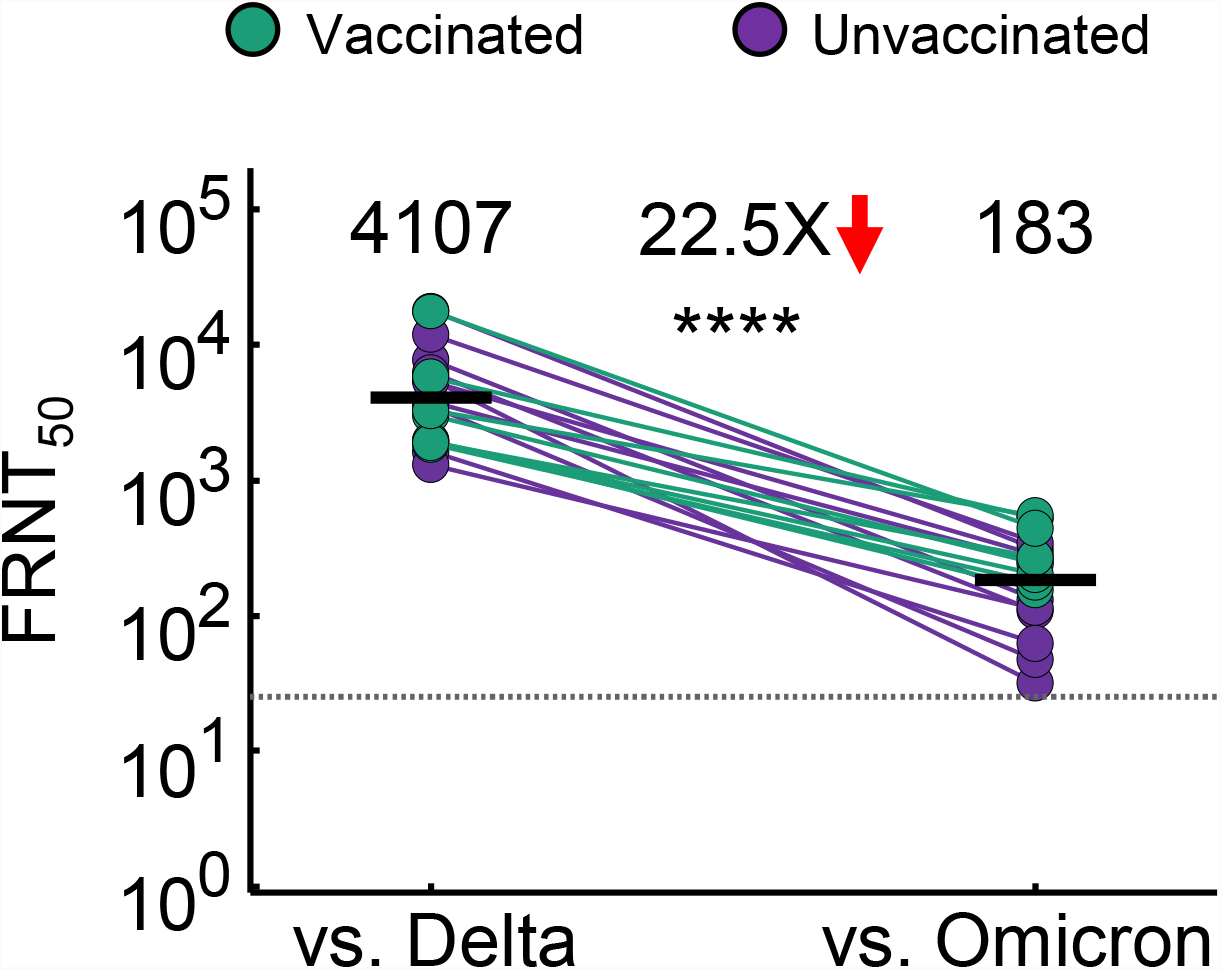
Escape of Omicron virus from Delta infection elicited immunity. Neutralization of Delta compared to Omicron virus by Delta infection elicited plasma immunity in vaccinated and unvaccinated participants. 18 samples were tested from n=14 participants infected during the Delta infection wave in South Africa (see Table S4). Dashed line is the most concentrated plasma tested. p-value is 1.6 *×*10^*−*7^ as determined by the Wilcoxon rank sum test.

The variability in Delta virus neutralization which we observed in the responses of unvaccinated participants may be because of previous unreported infection in some individuals, which could potentially confer a degree of Delta immunity. In contrast, vaccinated participants all had previous SARS-CoV-2 immunity from vaccination. They showed a stronger rise in Omicron neutralization and stronger enhancement of immunity to the Delta variant relative to unvaccinated participants. The dependence of Delta neutralization enhancement on previous immunity may indicate that enhancement may rely on boosting previous SARS-CoV-2 immunity rather than elicit antibodies that can specifically recognize and neutralize both Omicron and Delta.

Vaccination leads to a lower hospitalization rate with Omicron infection (https://www.discovery.co.za/corporate/health-insights-vaccines-real-world-effectiveness). This may be because Omicron does not have extensive escape from other arms of the adaptive immune response^12^ or because Omicron virus shows attenuated cell-to-cell spread^13^ which leads to decreased lung infection and pathology^14,15^. A stronger neutralizing response after Omicron infection, as shown here in vaccinated participants, should also contribute to vaccine mediated protection against more severe disease with Omicron.

A limitation of this study is the heterogeneity in participant immune history. Among the 10 vaccinated, there were two vaccination types and one participant was boosted. Among the unvaccinated, it is likely that some had unreported previous infection, based on the high seroprevalence observed in South Africa^16,17^. This heterogeneity is reflective of the increasing complexity of the SARS-CoV-2 immunity landscape as more people are infected with different variants and vaccinated with different vaccines. Despite the heterogeneity, there was a clear increase in both Omicron and Delta neutralization in the vaccinated group. In contrast, the trend in unvaccinated participants for Delta immunity was less clear. The majority of our participants were hospitalized which may be perceived as a limitation since Omicron infection is generally mild^18^. However, hospital admission should not be equated with severe disease. None of the participants in this study had severe disease as defined by the WHO ordinal scale, which requires at a minimum administration of high-flow oxygen^19^.

Work by us^2^ and others^3-8^ shows that residual vaccine elicited neutralizing immunity Omicron remains when neutralization capacity of pre-Omicron strains is high. Here we investigate a different situation, where there is breakthrough Omicron infection in vaccinated participants. Omicron neutralization capacity in these breakthrough cases was low and similar to the unvaccinated close to the time of infection. This is consistent with the notion that the reason these breakthroughs occurred was because of the low Omicron neutralization levels, likely the product of the antibody immune response waning, as vaccination was about four and a half months before infection. Despite low initial neutralization levels similar to the unvaccinated group, the response to Omicron was stronger in vaccinated participants and increased 13.7-fold between the enrollment and follow-up visits. As expected, Delta neutralization was higher in the vaccinated, and this also showed a stronger increase.

Based on neutralization levels, the immunity elicited in vaccinated individuals by Omicron infection is more potent against Delta relative to Omicron by a factor of approximately 2. Neutralization elicited by Delta infection is also more potent against Delta relative to Omicron, by a factor of over 20-fold. This may mean that in areas with a high frequency of infections and high vaccination coverage, Omicron is more likely than Delta to re-infect individuals who were previously infected either by Delta or Omicron. It may therefore dominate infections at the expense of Delta. In contrast, because unvaccinated individuals infected with Omicron develop a poor neutralization response against Delta, what may happen in areas of low vaccination coverage is less clear.

## Materials and methods

### Informed consent and ethical statement

Blood samples were obtained after written informed consent from adults with PCR-confirmed SARS-CoV-2 infection who were enrolled in a prospective cohort study approved by the Biomedical Research Ethics Committee at the University of KwaZulu–Natal (reference BREC/00001275/2020). Use of residual swab sample for SARS-0CoV-2 isolation was approved by the University of the Witwatersrand Human Research Ethics Committee (HREC) (ref. M210752).

### Data availability statement

Sequence of outgrown virus has been deposited in GISAID with accession EPI_ISL_7886688. Raw images of the data are available upon reasonable request.

### Code availability

Curve fitting scripts in MATLAB v.2019b are available on GitHub (https://github.com/sigallab/NatureMarch2021).

### Whole-genome sequencing, genome assembly and phylogenetic analysis

RNA was extracted on an automated Chemagic 360 instrument, using the CMG-1049 kit (Perkin Elmer, Hamburg, Germany). The RNA was stored at −80°C prior to use. Libraries for whole genome sequencing were prepared using either the Oxford Nanopore Midnight protocol with Rapid Barcoding or the Illumina COVIDseq Assay. For the Illumina COVIDseq assay, the libraries were prepared according to the manufacturer’s protocol. Briefly, amplicons were tagmented, followed by indexing using the Nextera UD Indexes Set A. Sequencing libraries were pooled, normalized to 4 nM and denatured with 0.2 N sodium acetate. A 8 pM sample library was spiked with 1% PhiX (PhiX Control v3 adaptor-ligated library used as a control). We sequenced libraries on a 500-cycle v2 MiSeq Reagent Kit on the Illumina MiSeq instrument (Illumina). On the Illumina NextSeq 550 instrument, sequencing was performed using the Illumina COVIDSeq protocol (Illumina Inc, USA), an amplicon-based next-generation sequencing approach. The first strand synthesis was carried using random hexamers primers from Illumina and the synthesized cDNA underwent two separate multiplex PCR reactions. The pooled PCR amplified products were processed for tagmentation and adapter ligation using IDT for Illumina Nextera UD Indexes. Further enrichment and cleanup was performed as per protocols provided by the manufacturer (Illumina Inc). Pooled samples were quantified using Qubit 3.0 or 4.0 fluorometer (Invitrogen Inc.) using the Qubit dsDNA High Sensitivity assay according to manufacturer’s instructions. The fragment sizes were analyzed using TapeStation 4200 (Invitrogen). The pooled libraries were further normalized to 4nM concentration and 25 μL of each normalized pool containing unique index adapter sets were combined in a new tube. The final library pool was denatured and neutralized with 0.2N sodium hydroxide and 200 mM Tris-HCL (pH7), respectively. 1.5 pM sample library was spiked with 2% PhiX. Libraries were loaded onto a 300-cycle NextSeq 500/550 HighOutput Kit v2 and run on the Illumina NextSeq 550 instrument (Illumina, San Diego, CA, USA). For Oxford Nanopore sequencing, the Midnight primer kit was used as described by Freed and Silander55. cDNA synthesis was performed on the extracted RNA using LunaScript RT mastermix (New England BioLabs) followed by gene-specific multiplex PCR using the Midnight Primer pools which produce 1200bp amplicons which overlap to cover the 30-kb SARS-CoV-2 genome. Amplicons from each pool were pooled and used neat for barcoding with the Oxford Nanopore Rapid Barcoding kit as per the manufacturer’s protocol. Barcoded samples were pooled and bead-purified. After the bead clean-up, the library was loaded on a prepared R9.4.1 flow-cell. A GridION X5 or MinION sequencing run was initiated using MinKNOW software with the base-call setting switched off. We assembled paired-end and nanopore.fastq reads using Genome Detective 1.132 (https://www.genomedetective.com) which was updated for the accurate assembly and variant calling of tiled primer amplicon Illumina or Oxford Nanopore reads, and the Coronavirus Typing Tool56. For Illumina assembly, GATK HaploTypeCaller -- min-pruning 0 argument was added to increase mutation calling sensitivity near sequencing gaps. For Nanopore, low coverage regions with poor alignment quality (<85% variant homogeneity) near sequencing/amplicon ends were masked to be robust against primer drop-out experienced in the Spike gene, and the sensitivity for detecting short inserts using a region-local global alignment of reads, was increased. In addition, we also used the wf_artic (ARTIC SARS-CoV-2) pipeline as built using the nextflow workflow framework57. In some instances, mutations were confirmed visually with .bam files using Geneious software V2020.1.2 (Biomatters). The reference genome used throughout the assembly process was NC_045512.2 (numbering equivalent to MN908947.3). For lineage classification, we used the widespread dynamic lineage classification method from the ‘Phylogenetic Assignment of Named Global Outbreak Lineages’ (PANGOLIN) software suite (https://github.com/hCoV-2019/pangolin)19. P2 stock was sequenced and confirmed Omicron with the following substitutions: E:T9I,M:D3G,M:Q19E,M:A63T,N:P13L,N:R203K,N:G204R,ORF1a:K856R,ORF1a:L2084I,ORF1a:A2710T, ORF1a:T3255I,ORF1a:P3395H,ORF1a:I3758V,ORF1b:P314L,ORF1b:I1566V,ORF9b:P10S,S:A67V,S:T95I, S:Y145D,S:L212I,S:G339D,S:S371L,S:S373P,S:S375F,S:K417N,S:N440K,S:G446S,S:S477N,S:T478K,S:E4 84A,S:Q493R,S:G496S,S:Q498R,S:N501Y,S:Y505H,S:T547K,S:D614G,S:H655Y,S:N679K,S:P681H,S:N76 4K,S:D796Y,S:N856K,S:Q954H,S:N969K,S:L981F. Sequence was deposited in GISAID, accession: EPI_ISL_7886688.

### Cells

Vero E6 cells (ATCC CRL-1586, obtained from Cellonex in South Africa) were propagated in complete growth medium consisting of Dulbecco’s Modified Eagle Medium (DMEM) with 10% fetal bovine serum (Hyclone) containing 10mM of HEPES, 1mM sodium pyruvate, 2mM L-glutamine and 0.1mM nonessential amino acids (Sigma-Aldrich). Vero E6 cells were passaged every 3–4 days. H1299 cell lines were propagated in growth medium consisting of complete Roswell Park Memorial Institute (RPMI) 1640 medium with 10% fetal bovine serum containing 10mM of HEPES, 1mM sodium pyruvate, 2mM L-glutamine and 0.1mM nonessential amino acids. H1299 cells were passaged every second day. The H1299-E3 (H1299-ACE2, clone E3) cell line was derived from H1299 (CRL-5803) as described in our previous work^2,20^.

### Virus expansion

All work with live virus was performed in Biosafety Level 3 containment using protocols for SARS-CoV-2 approved by the Africa Health Research Institute Biosafety Committee. ACE2-expressing H1299-E3 cells were seeded at 4.5 × 10^5^ cells in a 6 well plate well and incubated for 18–20 h. After one DPBS wash, the sub-confluent cell monolayer was inoculated with 500 μL universal transport medium diluted 1:1 with growth medium filtered through a 0.45-μm filter. Cells were incubated for 1 h. Wells were then filled with 3 mL complete growth medium. After 4 days of infection (completion of passage 1 (P1)), cells were trypsinized, centrifuged at 300 rcf for 3 min and resuspended in 4 mL growth medium. Then all infected cells were added to Vero E6 cells that had been seeded at 2 × 10^5^ cells per mL, 20mL total, 18–20 h earlier in a T75 flask for cell-to-cell infection. The coculture of ACE2-expressing H1299-E3 and Vero E6 cells was incubated for 1 h and the flask was filled with 20 mL of complete growth medium and incubated for 4 days. The viral supernatant from this culture (passage 2 (P2) stock) was used for experiments.

### Live virus neutralization assay

H1299-E3 cells were plated in a 96-well plate (Corning) at 30,000 cells per well 1 day pre-infection. Plasma was separated from EDTA-anticoagulated blood by centrifugation at 500 rcf for 10 min and stored at −80 °C. Aliquots of plasma samples were heat-inactivated at 56 °C for 30 min and clarified by centrifugation at 10,000 rcf for 5 min. Virus stocks were used at approximately 50-100 focus-forming units per microwell and added to diluted plasma. Antibody–virus mixtures were incubated for 1 h at 37 °C, 5% CO_2_. Cells were infected with 100 μL of the virus–antibody mixtures for 1 h, then 100 μL of a 1X RPMI 1640 (Sigma-Aldrich, R6504), 1.5% carboxymethylcellulose (Sigma-Aldrich, C4888) overlay was added without removing the inoculum. Cells were fixed 18 h post-infection using 4% PFA (Sigma-Aldrich) for 20 min. Foci were stained with a rabbit anti-spike monoclonal antibody (BS-R2B12, GenScript A02058) at 0.5 μg/mL in a permeabilization buffer containing 0.1% saponin (Sigma-Aldrich), 0.1% BSA (Sigma-Aldrich) and 0.05% Tween-20 (Sigma-Aldrich) in PBS. Plates were incubated with primary antibody overnight at 4 °C, then washed with wash buffer containing 0.05% Tween-20 in PBS. Secondary goat anti-rabbit HRP conjugated antibody (Abcam ab205718) was added at 1 μg/mL and incubated for 2 h at room temperature with shaking. TrueBlue peroxidase substrate (SeraCare 5510-0030) was then added at 50 μL per well and incubated for 20 min at room temperature. Plates were imaged in an ImmunoSpot Ultra-V S6-02-6140 Analyzer ELISPOT instrument with BioSpot Professional built-in image analysis (C.T.L).

### Statistics and fitting

All statistics and fitting were performed using custom code in MATLAB v.2019b. Neutralization data were fit to:

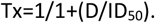

Here Tx is the number of foci normalized to the number of foci in the absence of plasma on the same plate at dilution D and ID_50_ is the plasma dilution giving 50% neutralization. FRNT_50_ = 1/ID_50_. Values of FRNT_50_ <1 are set to 1 (undiluted), the lowest measurable value. We note that the most concentrated plasma dilution was 1:25 and therefore FRNT_50_ < 25 were extrapolated.

## Data Availability

All data produced in the present work are contained in the manuscript or available upon reasonable request to the authors

## Acknowledgements

This study was supported by the Bill and Melinda Gates award INV-018944 (AS), National Institutes of Health award R01 AI138546 (AS), and South African Medical Research Council awards (AS, TdO, PLM) and the UK Foreign, Commonwealth and Development Office and Wellcome Trust (Grant no 221003/Z/20/Z, PLM). PLM is also supported by the South African Research Chairs Initiative of the Department of Science and Innovation and the NRF (Grant No 98341). The funders had no role in study design, data collection and analysis, decision to publish, or preparation of the manuscript.

---

**Table S1:** Read counts of majority and minority genotypes detected in outgrown virus used in experiments.

| Amino Acid change | Nucleotide change | Codon change | Reads |
| --- | --- | --- | --- |
| <u>A67V</u> | 21762C>T | 21761 GCT>GTT | GCT – 45 GTT – 3134 |
| <u>*H69_V70del</u> | 21766_21771delACATGT | 21766_21771ACATGT>del | ACATGT – 0 del – 1132 |
| <u>T95I</u> | 21846C>T | 21845 ACT>ATT | ACT – 53 ATT – 2171 |
| <u>*G142D</u> | 21987_21989delGTG | 21987_21989GTG >del | GTG – 0 del – 1572 |
| <u>*V143_Y145del</u> | 21990_21995delTTTATT | 21990_21995TTTATT >del | TTTATT – 0 del – 1572 |
| <u>*L212I</u> | 22194_22196delATT | 22194_22196ATT >del | ATT – 114 del – 2897 |
| <u>*R214_D215</u> | 22204_22205insGAGCCAGAA | 22204_22205GAGCCAGAA >ins | WT – 808 insGAGCCAGAA – 1316 |
| <u>G339D</u> | 22578G>A | 22577 GGT>GAT | GGT – 104 GAT – 3519 |
| <u>S371L</u> | 22674C>T | 22674 TCC>CTC | TCC – 41 CTC – 1359 |
| <u>S373P</u> | 22679T>C | 22679 TCA>CCA | TCA – 64 CCA – 1696 |
| <u>S375F</u> | 22686C>T | 22685 TCC>TTC | TCC – 25 TTC – 1569 |
| <u>K417N</u> | 22813G>T | 22811 AAG>AAT | AAG – 36 AAT – 1885 |
| <u>N440K</u> | 22882T>G | 22880 AAT>AAG | AAT – 294 AAG – 1579 |
| <u>G446S</u> | 22898G>A | 22898 GGT>AGT | GGT – 74 AGT – 1686 |
| <u>S477N</u> | 22992G>A | 22991 AGC>AAC | AGC – 18 AAC – 1917 |
| <u>T478K</u> | 22995C>A | 22994 ACA>AAA | ACA – 55 AAA – 1968 |
| <u>E484A</u> | 23013A>C | 23012 GAA>GCA | GAA – 32 GCA – 1903 |
| <u>Q493R</u> | 23040A>G | 23039 CAA>CGA | CAA – 4 CGA – 2060 |
| <u>G496S</u> | 23048G>A | 23048 GGT>AGT | GGT – 53 AGT – 1734 |
| <u>Q498R</u> | 23055A>G | 23054 CAA>CGA | CAA – 28 CGA – 1733 |
| <u>N501Y</u> | 23063A>T | 23063 AAT>TAT | AAT – 49 TAT – 1812 |
| <u>Y505H</u> | 23075T>C | 23075 TAC>CAC | TAC – 55 CAC – 1451 |
| <u>T547K</u> | 23202C>A | 23201 ACA>AAA | ACA – 10 AAA – 1655 |
| <u>D614G</u> | 23403A>G | 23402 GAT>GGT | GAT – 17 GGT – 1398 |
| <u>H655Y</u> | 23525C>T | 23525 CAT>TAT | CAT – 26 TAT – 1556 |
| <u>N679K</u> | 23599T>G | 23597 AAT>AAG | AAT – 21 AAG – 1245 |
| <u>P681H</u> | 23604C>A | 23603 CCT>CAT | CCT – 0 CAT – 535 |
| <u>N764K</u> | 23854C>A | 23852 AAC>AAA | AAC – 23 AAA – 290 |
| <u>D796Y</u> | 23948G>T | 23948 GAT>TAT | GAT – 8 TAT – 217 |
| <u>N856K</u> | 24130C>A | 24128 AAC>AAA | AAC – 9 AAA – 155 |
| <u>Q954H</u> | 24424A>T | 24422 CAA>CAT | CAA – 9 CAT – 298 |
| <u>N969K</u> | 24469T>A | 24467 AAT>AAA | AAT – 31 AAA – 392 |
| <u>L981F</u> | 24503C>T | 24503 CTT>TTT | CTT – 112 TTT – 347 |

\*Only deletions or insertions where the adjacent codon was preserved were counted. WT – Wild Type i.e reads without the insertion.

**Table S2:** Summary characteristics of Omicron infected participants.

|  | All | Vaccinated | Unvaccinated |
| --- | --- | --- | --- |
| Number Participants | 23 | 10 | 13 |
| Age* | 32 (26-37) | 36 (33-36) | 26 (26-34) |
| Male sex | 10 (43%) | 4 (40%) | 6 (46%) |
| Days post-vaccination |  | 140 (116-192) |  |
| Days post-symptom onset to enrolment | 5 (3-8) | 4 (2-6) | 7 (3-8) |
| Days post-symptom onset to last follow-up | 23 (17-25) | 23 (15-27) | 22 (19-24) |

\*Median (IQR).

**Table S3:** Detailed characteristics of Omicron infected participants.

| Participant # | Age | Sex | Vacc. type | Vacc. date | Days post-vacc. to enroll. | Date symptom onset | Ct enroll. | Symptoms onset to last follow-up | GISAID ID of infecting virus |
| --- | --- | --- | --- | --- | --- | --- | --- | --- | --- |
| 1 | 30-39 | M | AD26.COV | Mar-2021 | 278 | Dec-2021* | 25 | 23 |  |
| 2 | 30-39 | M | AD26.COV** | Mar-2021 | 264 | Nov-2021 | 14 | 22 |  |
| 3 | 50-59 | F | BNT162b2 | May-2021 | 200 | Dec-2021 | 17 | 27 | EPI_ISL_8604915 |
| 4 | 30-39 | F | AD26.COV | May-2021 | 210 | Dec-2021 | 31 | 13 | EPI_ISL_8604910 |
| 5 | 20-29 | F | AD26.COV | Sep-2021 | 89 | Dec-2021 | 24 | 27 |  |
| 6 | 10-19 | F | BNT162b2 | Jul-2021 | 157 | Dec-2021 | 23 | 12 | EPI_ISL_8604906 |
| 7 | 20-29 | F | No |  |  | Nov-2021 | UND | 24 |  |
| 8 | 30-39 | M | No |  |  | Dec-2021 | 18 | 23 | EPI_ISL_8604919 |
| 9 | 40-49 | F | No |  |  | Dec-2021 | 32 | 28 | EPI_ISL_8604901 |
| 10 | 20-29 | M | No |  |  | Dec-2021 | 30 | 13 | EPI_ISL_8604908 |
| 11 | 20-29 | F | No |  |  | Dec-2021 | 28 | 22 | EPI_ISL_8604913 |
| 12 | 20-29 | F | No |  |  | Dec-2021* | UND | 22 |  |
| 13 | 30-39 | M | BNT162b2 | Jul-2021 | 129 | Nov-2021 | 32 | 28 | EPI_ISL_8604916 |
| 14 | 20-29 | M | No |  |  | Nov-2021 | 31 | 15 |  |
| 15 | 60-69 | F | BNT162b2 | May-2021 | 198 | Dec-2021 | 25 | 25 | EPI_ISL_8604920 |
| 16 | 60-69 | M | BNT162b2 | Dec-2021 | 15 | Dec-2021 | 24 | 10 | EPI_ISL_8578311 |
| 17 | 30-39 | M | No |  |  | Dec-2021 | 37 | 19 | EPI_ISL_8604923 |
| 18 | 60-69 | F | No |  |  | Dec-2021<sup>#</sup> | 27 | 23 | EPI_ISL_8578312 |
| 19 | 30-39 | M | No |  |  | Dec-2021* | 31 | 27 | EPI_ISL_8604924 |
| 20 | 20-29 | F | No |  |  | Dec-2021 | 37 | 24 | EPI_ISL_8604911 |
| 21 | 20-29 | M | No |  |  | Dec-2021 | 28 | 21 | EPI_ISL_8604922 |
| 22 | 30-39 | F | AD26.COV | Aug-2021 | 120 | Dec-2021 | 33 | 23 |  |
| 23 | 20-29 | F | No |  |  | Dec-2021 | 30 | 10 | EPI_ISL_8604902 |

Ct enroll.: qPCR cycle threshold for SARS-CoV-2 at enrollment. UND: Undetectable. AD26.COV: Johnson and Johnson AD26.CoV.2 vaccine. \*Reported previous infection. \*\*Boosted with Ad26.CoV2.S in Nov-2021.<sup>#</sup>Required supplemental O<sub>2</sub>.

**Table S4:**
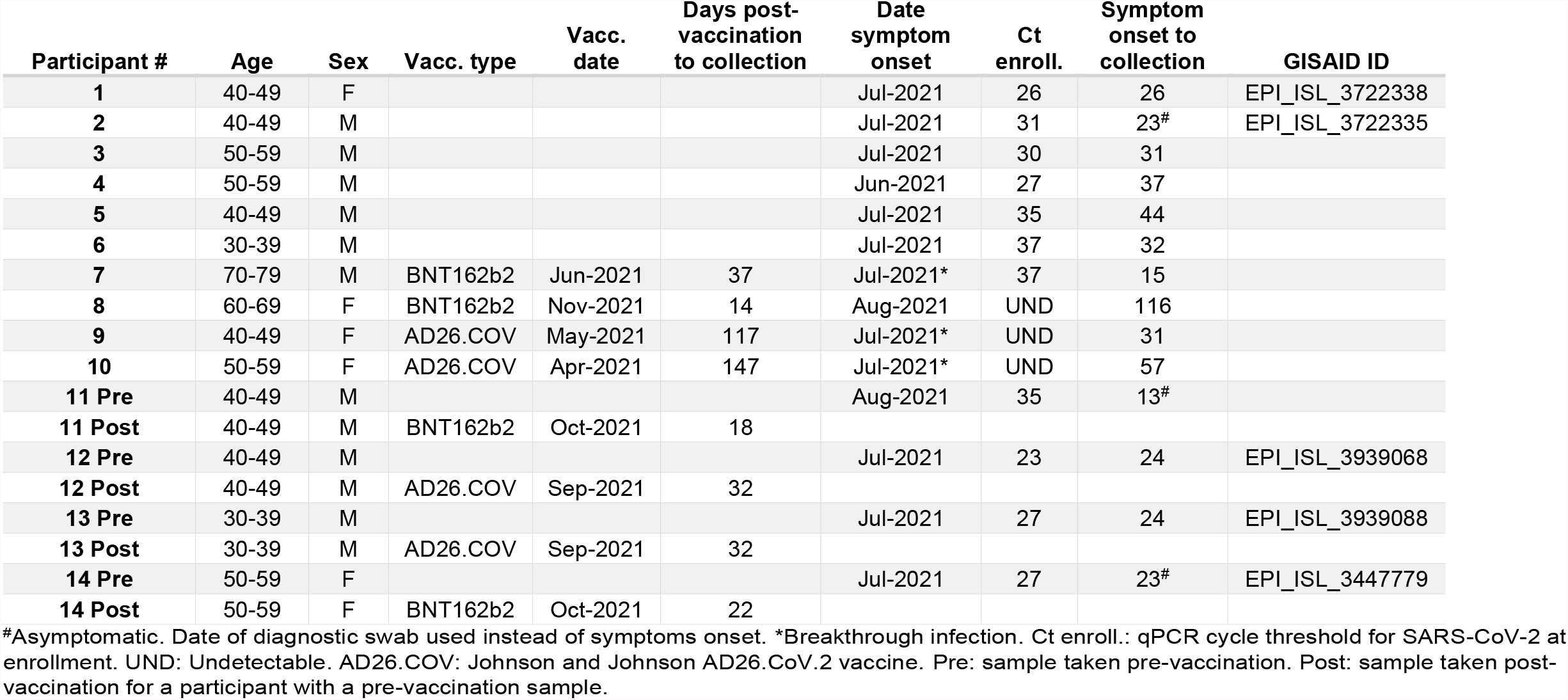
Detailed characteristics of Delta infected participants.

| Participant # | Age | Sex | Vacc. type | Vacc. date | Days post-vaccination to collection | Date symptom onset | Ct enroll. | Symptom onset to collection | GISAID ID |
| --- | --- | --- | --- | --- | --- | --- | --- | --- | --- |
| 1 | 40-49 | F |  |  |  | Jul-2021 | 26 | 26 | EPI_ISL_3722338 |
| 2 | 40-49 | M |  |  |  | Jul-2021 | 31 | 23<sup>#</sup> | EPI_ISL_3722335 |
| 3 | 50-59 | M |  |  |  | Jul-2021 | 30 | 31 |  |
| 4 | 50-59 | M |  |  |  | Jun-2021 | 27 | 37 |  |
| 5 | 40-49 | M |  |  |  | Jul-2021 | 35 | 44 |  |
| 6 | 30-39 | M |  |  |  | Jul-2021 | 37 | 32 |  |
| 7 | 70-79 | M | BNT162b2 | Jun-2021 | 37 | Jul-2021* | 37 | 15 |  |
| 8 | 60-69 | F | BNT162b2 | Nov-2021 | 14 | Aug-2021 | UND | 116 |  |
| 9 | 40-49 | F | AD26.COV | May-2021 | 117 | Jul-2021* | UND | 31 |  |
| 10 | 50-59 | F | AD26.COV | Apr-2021 | 147 | Jul-2021* | UND | 57 |  |
| 11 Pre | 40-49 | M |  |  |  | Aug-2021 | 35 | 13<sup>#</sup> |  |
| 11 Post | 40-49 | M | BNT162b2 | Oct-2021 | 18 |  |  |  |  |
| 12 Pre | 40-49 | M |  |  |  | Jul-2021 | 23 | 24 | EPI_ISL_3939068 |
| 12 Post | 40-49 | M | AD26.COV | Sep-2021 | 32 |  |  |  |  |
| 13 Pre | 30-39 | M |  |  |  | Jul-2021 | 27 | 24 | EPI_ISL_3939088 |
| 13 Post | 30-39 | M | AD26.COV | Sep-2021 | 32 |  |  |  |  |
| 14 Pre | 50-59 | F |  |  |  | Jul-2021 | 27 | 23<sup>#</sup> | EPI_ISL_3447779 |
| 14 Post | 50-59 | F | BNT162b2 | Oct-2021 | 22 |  |  |  |  |

<sup>#</sup>Asymptomatic. Date of diagnostic swab used instead of symptoms onset. \*Breakthrough infection. Ct enroll.: qPCR cycle threshold for SARS-CoV-2 at enrollment. UND: Undetectable. AD26.COV: Johnson and Johnson AD26.CoV.2 vaccine. Pre: sample taken pre-vaccination. Post: sample taken post-vaccination for a participant with a pre-vaccination sample.

## Notes

### Competing Interest Statement

The authors have declared no competing interest.

### Author Declarations

Blood samples were obtained after written informed consent from adults with PCR-confirmed SARS-CoV-2 infection who were enrolled in a prospective cohort study approved by the Biomedical Research Ethics Committee at the University of KwaZulu-Natal (reference BREC/00001275/2020). Use of residual swab sample was approved by the University of the Witwatersrand Human Research Ethics Committee (HREC) (ref. M210752).

### Summary of Updates

Added more participants to the study.

